# Hsa-miRNA-1278 is a Novel Predictor for the Hematoma Expansion of Intracerebral Hemorrhage

**DOI:** 10.1101/2023.05.16.23290076

**Authors:** Na Li, Kaijiang Kang, Zeqiang Ji, Feng Zhang, Xingquan Zhao

## Abstract

**Background and Purpose:** Hematoma expansion has been identified as a crucial predictor for the progress and outcome of intracranial hemorrhage(ICH). MicroRNAs (miRNAs) are associated with hematoma expansion and play important roles in regulating the mechanism of ICH. Here, we identified a miRNA, hsa-miR-1278, as a predictor of hematoma expansion and earlier estimation of ICH prognosis.

**Methods:** The participants who had been diagnosed with ICH by brain imaging were divided into hematoma enlargement(HE) group and non-HE group. A total of 10 samples from 2 groups were extracted and subjected to high-throughput sequencing. The differentially expressed microRNAs (DEMs) were identified by bioinformatics and quantitated by reverse transcription-polymerase quantitive chain reaction(RT-qPCR). To further validate the DEMs, We searched the Targescan database to find the target gene of the DEM and cultured the QSG7701 cells line and performed western blotting to validated the target miRNA. The Continuous variables were expressed as mean ± SD and analyzed by unpaired Student’s t-test; categorical variables were analyzed by chi-square test, and P values < 0.05 were considered statistically significant.

**Results:** We performed miRNA sequencing of HE and non-HE in 10 patients with cerebral hemorrhage. We found 18 differentially expressed miRNAs in HE group. We then performed RT-qPCR verification and identified that hsa-miR-1278 was significantly increased in the HE group (P <0.05). By searching Targescan database and genes related to ICH, we selected IL22 and PF4 as the target genes of has-miR-1278. RT-qPCR showed that PF4 were decrease in HE, which was consistent with the increased of hsa-miR-1278.

**Conclusions:** The expression of hsa-miR-1278 was still significantly up-regulated in the hematoma expansion group, and therefore made the hsa-miR-1278 as a novel predictor of ICH prognosis.

## Introduction

Intracerebral hemorrhage (ICH) is the most devastating form of stroke due to the highest disability and the mortality rates(1, 2). Hematoma volume is a crucial predictor of 30-day mortality after ICH(3), and hematoma expansion (HE) has been identified as one of the most important independent determinants of early neurological deterioration and poor clinical outcome after ICH(4, 5). Although hemostatic therapy and intensive blood pressure reduction treatment can reduce the growth of hematoma, both of them failed to improve clinical outcome after ICH(6–9) Accurate identification of patients with high risk of hematoma growth is crucial for developing anti-expansion therapies.

The computed tomographic angiography (CTA) contrast extravasation, also called spot sign, is a radiological technology for predicting HE and poor clinical outcome(10, 11). Clinical predictive score using predictors including CTA spot sign has been developed(12) However, previous study showed poor calibration of CTA(13), and it is not available in routine diagnostic workup of acute ICH in many institutions. Moreover, spot sign has a lower-than-expected prevalence rate as well as low sensitivity to predict patients at risk of hematoma expansion(14). Recently, several non-contrast computed tomography (NCCT) imaging predictors of HE have been developed, such as hypodensity, density heterogeneity, swirl sign, black hole sign, blend sign, margin irregularity and island sign. Some of them showed overlapping radiological messages(15–21). A 5-point BAT score including swirl sign, hypodensity and timing of NCCT has also been established(22). However, the main deficit is the low sensitivity, with more than half of the expanders failed to be detected, resulting in a high rate of error prediction(23). Therefore, it is crucial to develop novel models for accurately predicting HE, ideally obtainable and operable from routine clinical NCCT.

In recent years, several studies have successfully proved the presence of miRNAs in human circulatory system, which are released from tissue cells, and may associate with diseases(24, 25). Because MicroRNAs are non-coding single-stranded RNAs containing 21–23 nucleotides that control the activity of various protein-coding genes through post-transcriptional repression. Previous studies have shown that circulating miR-26a could act as a potential predictor and therapeutic target for non-hypertensive ICH(26); miR-146a is significantly down-regulated in ICH rats, inhibiting inflammation and oxidative stress to protect against intracerebral hemorrhage(27); and miR-139 is activated to protect cerebral hemorrhage injury in rats(28). Therefore, miRNAs as the novel biomarkers for the early diagnosis of disease would be a promising strategy.

In the present study, we detected the expression pattern of miRNAs in peripheral blood samples from HE and non-HE patients, and identified hsa-miR-1278 as a novel biological marker for ICH prognosis. Subsequent experiments including real time-quantitative PCR (RT-qPCR) and western blot assays further validated the significance of the discovery.

## Materials and Methods

### Clinical specimens and study design

We were collected between Aug 2017 and Jul 2019, from 132 patients treated at Beijing Tiantan Hospital, Capital Medical University who met the following characteristics: primary spontaneous supratentorial hemorrhages with baseline volume ≤60 mL, presentation to the emergency department within 6 hours from symptom onset, lack of anticoagulant treatment, and international normalized ratio <1.5. The exclusion criteria: traumatic intracranial bleeding, tumor or vascular malformation underlying the hemorrhage, hemorrhagic conversion of ischemic stroke, primary intraventricular hemorrhage, and missing follow-up NCCT. HE is defined as relative hematoma growth >33% or absolute hematoma growth >6 mL from baseline hemorrhage volume. The study was performed according to the guidelines from the Helsinki Declaration, and was approved by the Research Ethics Committee of the hospital. Written informed consent was obtained from all the subjects or their legally authorized representatives.

### miRNA sequencing

ICH patients were divided into two groups. The HE group includes stroke patients with enlarged hematoma, while the non-bleeding patients are non-HE group. Total RNA of blood samples from different groups were extracted using Qiagene kit (Cat# 52304) according to the manufacturer’s protocol. RNA samples were quantified using a UV spectrophotometry (NanoDrop 2000, Thermo Scientific). Quality of total RNA was evaluated using total RNA Pico chip analysis on Agilent 2100 Bioanalyzer. MiRNA libraries from ribosomal RNA (rRNA) depleted total RNA (Ribo-Zero rRNA removal Kit, Epicentre) were subjected to miRNA sequencing on a BGISEQ-500 platform. (BGI-Shenzhen, China) The sample reads were trimmed to remove reads with an unknown base (N) content greater than 5%, adapters and low-quality bases using Trimmomatic software and aligned with the reference genome using HISAT and Bowtie2 software.

### The identification of differentially expressed miRNA

The inspection standard of differential expression miRNA was listed below:

1. For the single replicates, miRNA with a large number of reads (both samples greater than 20) were tested by the χ2 test, and the less were tested by Fisher’s extract test;
2. For multiple replicates, the permutation test was used without presupposing its distribution.

The P value obtained by the statistical test is corrected by false discovery rate (FDR) to calculate the Q value.

Differential miRNA expression was identified when

1. Log_2_(fold change)>1 or log_2_(fold change)<-1
2. P<0.05 (significant difference)
3. Average expression in at least one group>1 (medium or high expression)

### RT-qPCR

The synthesis of cDNA were performed by using miRcute Plus miRNA First-Strand cDNA Synthesis Kit (TIANGEN) according to the manufacturer’s instructions. For quantitative assessment of targeted miRNA, reverse transcribed RNA were measured by miRcute Plus miRNA qPCR Detection Kit (TIANGEN). TaqMAN probe synthesis from Shanghai Biotech Biotechnology Co. We designed specific primers for the detection sequences: hsa-miR-1278:5’CGCGCGTAGTACTGTGCATATC 3’,5’AGTGCAGGGTCCGAG GTATT 3’;hsa-miR-1303:5’ CGCGTTTAGAGACGGGGTCT 3’,5’AGTGCAGGG TCCGAGGTATT 3’;hsa-miR-1227:5’GCGCGGTGGGGCCAGG 3’,5’ AGTGCAG GGTCCGAGGTATT 3’;IL-22:L:TCCTACCACCAGGGCGATTA,R:ATCGGGGT TGTCTGCTCTTG;PF4:L:AGCCACACTTAACGGAGAGC; R:CACACACG TAGGCAGCTAGT.

Cycle threshold(Ct) values of the analyzed miRNAs were normalized to Ct values obtained for the noncoding, small nuclear RNA molecule U6. RT-qPCR was performed using an ABI 7500 Fluorescent Quantitative PCR System (Applied Biosystems, USA). Expression changes were calculated using 2^−ΔΔCt^ methods. All statistical analyses were performed at least three times in independent experiments and analyzed using two-tailed unpaired Student’s t-tests. P≦0.05 was considered as statistically significant.

### Cell culture and miRNA silencing

QSG7701 (Sciencell Research Laboratories,Inc,USA) cells were cultured using RPMI medium (Gibco) containing 10% fetal bovine serum (Gibco) in a 37℃, 5% CO_2_ constant temperature cell incubator.Hsa-miR-1278 silencer (Thermo Fisher) and negative controls were transfected into QSG7701 cells. After 24 h of transfection, Total RNA was extracted using a Qiagene Kit (Cat# 72405). The RNA concentration was measured using a Nanodrop 2.0 nucleic acid analyzer. RNA samples with an absorbance (A260/280 nm) ratio of 1.8 to 2.0 were taken and reversed to transcribe into cDNA according to the reverse transcription kit instructions. The expression of hsa-miR-1278 were detected using TanMan fluorescent quantitative PCR probes, and small nuclear RNA (U6) was used as an internal reference. RT-qPCR reaction condition was as the following: TanMan Mix II 10µL, cDNA 1µL, TanMan miR-26b primer probe 1 µL, sterilized ddH2O 8 µL. Cycle parameters: 95 ℃ for 10 min; 95 ℃ for 15s, 60 ℃ for 1 min, a total of 40 cycles. The relative expression of miRNAs was calculated by 2^-△△ Ct^ method.

### Protein blot analysis

QSG7701 cells were collected and lysed in eukaryotic cell lysis buffer (BioDev-Tech., Beijing, China) according to the manufacturer’s instructions. Proteins (100 µg) were mixed and boiled in SDS-PAGE sample buffer for 5 min, separated by SDS-PAGE (12% polyacrylamide gel), and then electro-transferred to nitrocellulose membranes by electroblotting in a Mini-Trans-Blot. The transferred membranes were washed for 10 min with TTBS buffer containing (in mM): 10 TrisHCl, pH 7.4, 150 NaCl and 0.05% (w/v) Tween-20, followed by a 5% skim milk in TTBS in a closed solution for 1 hr. The closed membranes were incubated with primary purified goat polyclonal antibodies against Kir6.1 and Kir6.2 (Santa Cruz Biotechnology, CA, USA) at a 1:2000 dilution for 3 h at room temperature. For secondary antibody, the fluorescently labeled secondary antibody (IRDye 680 donkey anti-rabbit IgG) was diluted in TTBS buffer and incubated for 1 h at room temperature. Membranes were scanned at 680 nm and 780 nm with an Odyssey infrared imaging system (Li COR, Biosciences, Lincoln, NE)

### Statistical analysis

Continuous variables were presented as mean±SD (normal distribution data) or median with interquartile range (skew distribution data), as appropriate. Categorical variables were presented as numbers and percentages. The χ2 test (categorical variables), t-test (normal distribution) and logistic regression were used to analyze the differences in characteristics between the HE and non-HE group. Variables with P < 0.05 in univariate analysis were included for multivariate analysis. The statistical analysis was performed using a commercial statistical software package (SPSS for Windows, version 22.0, IBM-SPSS, Chicago, IL, US).

## Results

### Patient characteristics

Between August 2017 and July 2019, 132 with ICH treated at Beijing Tiantan Hospital, Capital Medical University were enrolled in this study. The cohort was divided into HE group and non-HE group based on the 24 ±3h follow-up CT. We analyzed the differences in clinical characteristics between the HE group and non-HE group by univariate analysis, which showed significant differences between the two groups in hsa-miRNA-1278, gender, history of hypertension, baseline SBP, and baseline ALT(p<0.05) The detailed characteristics were shown in Table 1.

**Table 1.** Baseline characteristics between patients with or without hematoma expansion (n=86)

| Variables | HE<br>(n=29) | Non-HE<br>(n=57) | *P* value |
| --- | --- | --- | --- |
| Male, n (%) | 26 (89.7%) | 40 (70.2%) | 0.043 |
| Age, mean ±SD | 52.9 ± 11.1 | 54.5 ± 11.6 | 0.531 |
| History of hypertension | 25 (92.6%) | 39 (70.9%) | 0.026 |
| SBP, mean ±SD | 167.9 ± 24.2 | 180.5 ± 27.0 | 0.037 |
| DBP, mean ±SD | 101.8 ± 16.6 | 106.4 ± 19.7 | 0.283 |
| Hematoma volume, mean ±SD | 34.9 ± 27.5 | 28.8 ± 21.6 | 0.341 |
| Basal ganglia hematoma | 23 (100.0%) | 41 (87.2%) | 0.168 |
| WBC, mean ±SD | 9.9 ± 3.7 | 9.3 ± 3.6 | 0.477 |
| PLT, mean ±SD | 209.4 ± 59.1 | 232.2 ± 49.0 | 0.063 |
| INR, mean ±SD | 0.98 ± 0.23 | 0.92 ± 0.06 | 0.051 |
| Fbg, mean ±SD | 2.81 ± 1.66 | 2.60 ± 0.60 | 0.400 |
| APTT, mean ±SD | 27.0 ± 3.9 | 26.0 ± 4.2 | 0.316 |
| GLU, mean ±SD | 8.0 ± 3.5 | 8.7 ± 6.4 | 0.554 |
| Cr, mean ±SD | 106.0 ± 205.5 | 70.9 ± 48.0 | 0.222 |
| BUN, mean ±SD | 6.7 ± 6.7 | 5.1 ± 1.1 | 0.085 |
| TC, mean ±SD | 4.4 ± 0.8 | 4.5 ± 1.0 | 0.770 |
| TG, mean ±SD | 1.3 ± 0.6 | 1.2 ± 0.5 | 0.692 |
| HDL, mean ±SD | 1.2 ± 0.2 | 1.3 ± 0.3 | 0.666 |
| LDL, mean ±SD | 2.9 ± 0.5 | 2.7 ± 0.7 | 0.495 |
| GHb, mean ±SD | 5.5 ± 0.6 | 5.7 ± 1.0 | 0.530 |
| ALT, mean ±SD | 40.6 ± 32.1 | 29.7 ± 15.7 | 0.045 |
| AST, mean ±SD | 29.4 ± 19.0 | 23.7 ± 7.5 | 0.061 |
| ALP, mean ±SD | 77.6 ± 24.4 | 81.7 ± 18.6 | 0.579 |
| Hcy, mean ±SD | 13.9 ± 4.7 | 12.0 ± 5.1 | 0.447 |
| HsCRP, mean ±SD | 4.5 ± 4.3 | 5.4 ± 5.0 | 0.630 |
| hsa-miRNA-1227 | 27.6 ± 1.2 | 27.7 ± 1.3 | 0.716 |
| hsa-miRNA-1278 | 33.0 ± 0.6 | 33.3 ± 0.3 | 0.004 |
| hsa-miRNA-1303 | 21.7 ± 0.8 | 21.8 ± 0.8 | 0.554 |

**ICH, intracerebral hemorrhage; HE, hematoma expansion; SD, standard deviation; SBP, systolic blood pressure; DBP diastolic blood pressure; WBC, white blood cell; PLT, platelet; INR, international normalized ratio; Fbg, fibrinogen; APTT, activated partial thromboplastin time; GLU, glucose; Cr, creatinine; BUN, blood urea nitrogen; TC, total cholesterol; TG, Triglyceride; HDL, high-density lipoprotein; LDL, low-density lipoprotein; GHb, Glycosylated Hemoglobin; ALT, alanine aminotransferase; AST, aspartate transaminase ALP, alkaline phosphatase; Hcy, homocysteine; HsCRP, hypersensitive-c-reactive-protein; ICH expansion was defined as hematoma growth>33% or >6 mL.**

### Differentially expressed microRNAs

The peripheral blood of 10 subjects was subjected to microRNA sequencing using high-throughput sequencing, differentially expressed microRNAs (DEMs) were identified by bioinformatics. A total of 10 samples were obtained with miRNA sequences of Q20% > 99.4% and Q30% > 97.4%. A total of 18 differential miRNAs were obtained by bioinformatic analysis of miRNAs, of which 10 were up-regulated and 8 were down-regulated in the HE group compared to the non-HE group. (Figure 1). To further validate the accuracy of the DEMs, we expanded the clinical sample. 132 samples were selected, including 45 samples in the HE group 87 samples in the non-HE group. After sample processing and quality control 86 samples(HE=29,non-HE=57) were tested by RT-PCR and 46 samples showed degradation and could not be tested. The validation results showed that 15 miRNAs did not match the first sequencing results after the expanded sample validation, and 3 miRNAs were consistent with each other. Three miRNAs with consistent results were hsa-miR-1278, hsa-miR-1303 and hsa-miR-1227, of which hsa-miR-1278 expression was significantly upregulated in HE group (P<0.05) (Figure 2).

**Figure 1.**
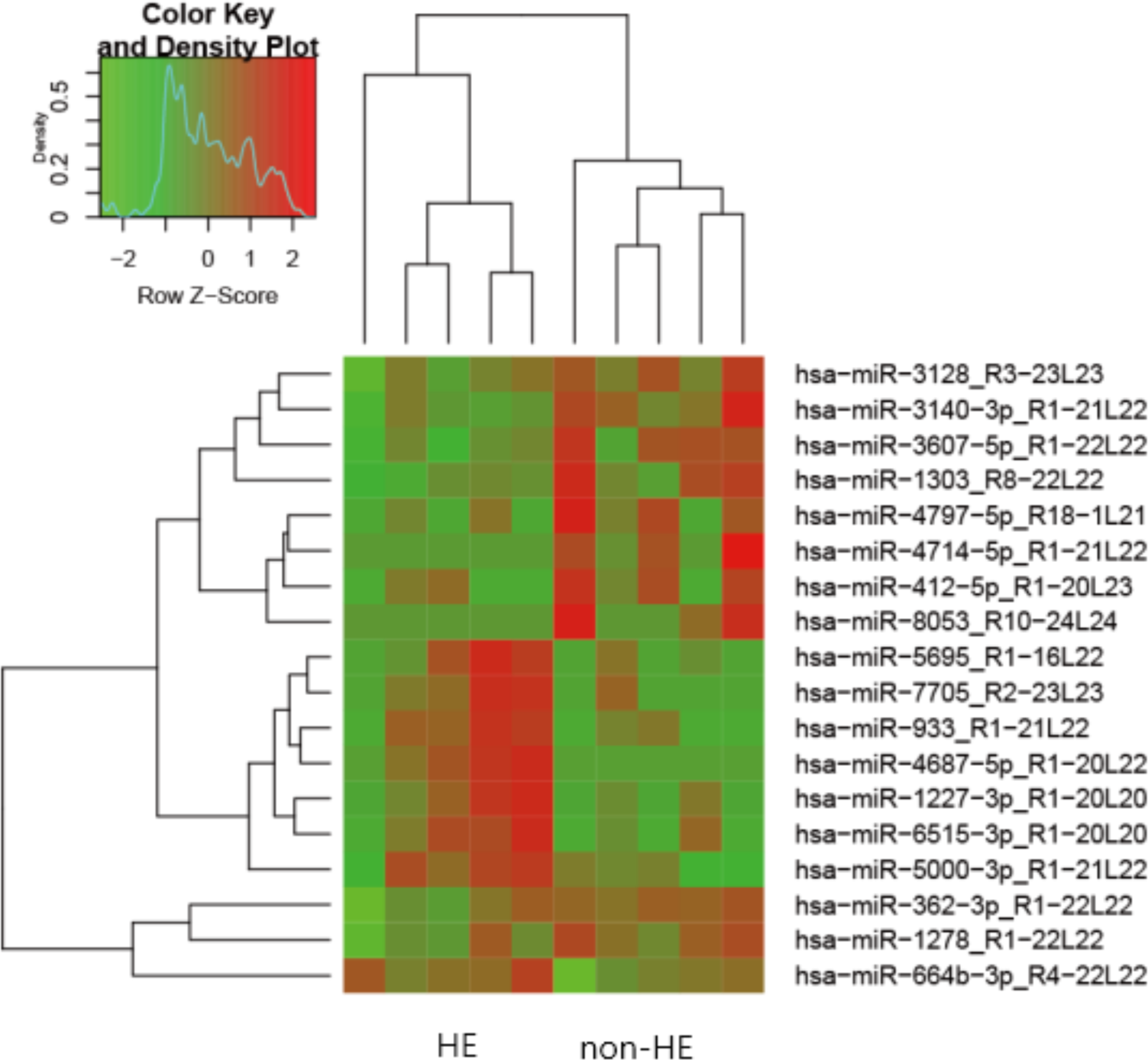
Heat map of differentially expressed miRNAs. The figure shows a two-way clustering using differentially expressed miRNAs and groups. The shades of color represent the level of miRNA expression, red represents high expression, and green represents low expression.

**Figure 2.**
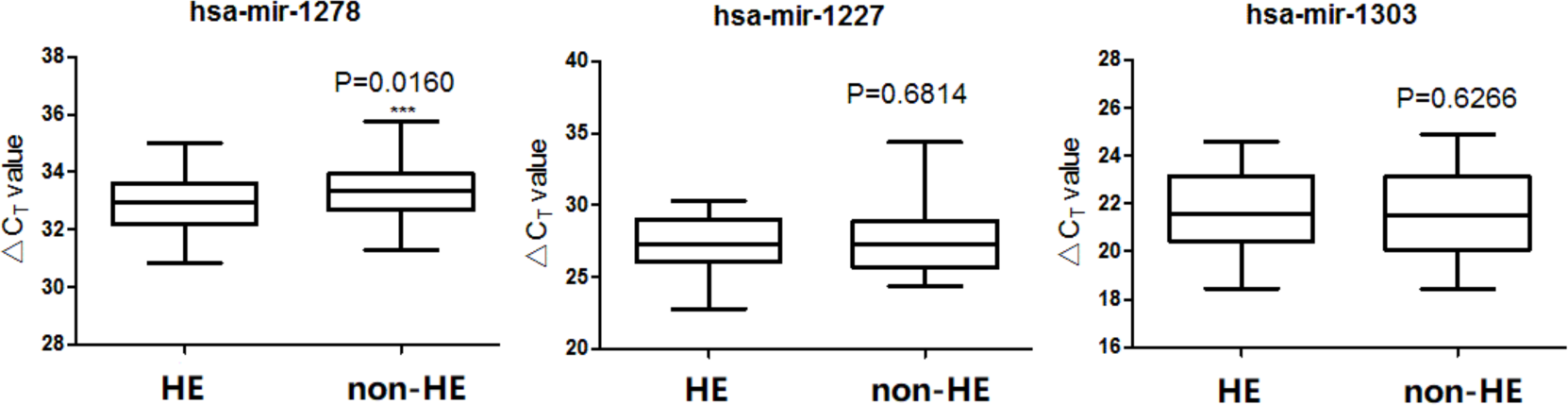
The differential expression miRNAs for RT-qPCR validation. The hsa-mir-1278 was down-regulated in HE group. P<0.05 was considered statistically significant.

### Identification of putative miRNA gene targets

We used miRNA target prediction database, Targetscan (http://www.targetscan.org) to identify potential miRNA-mRNA target pairs. After filtering target pairs including differentially expressed genes and miRNAs, we obtained a total of 1,922 miRNA-mRNA pairs, represented 309 genes and 174 known mature human miRNAs. Based on the screening of hsa-miR-1278 combined with cerebral hemorrhage symptoms and Targescan database, we identified IL22 and PF4 as candidate targets. We then performed RT-qPCR to verify the expression of IL22 and PF4 in HE and non-HE samples, of which the expression of PF4 gene was significantly downregulated in HE group. (P<0.05) (Figure 3)

**Figure 3.**
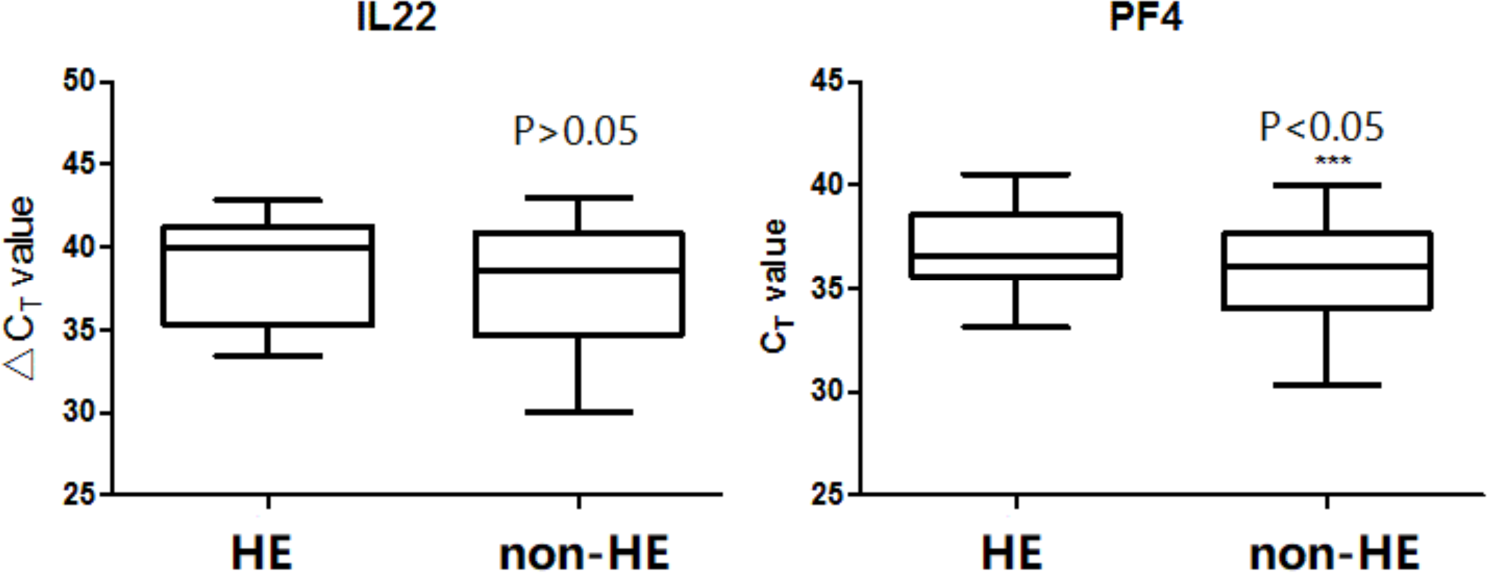
RT-qPCR validation of IL22 and PF4 in HE and non-HE groups. The expression of PF4 was considered to be down-regulated in HE group. ICH expansion was defined as hematoma growth>33% or >6 mL. P<0.05 was considered statistically significant.

### miRNA regulates target genes in cellular models

To further verify the protein targets of hsa-miR-1278, we performed inhibition experiment and western blotting using QSG-7701 cells (Human hepatocyte line). We first carried out miRNA silencing experiments in QSG-7701 cells. Morphology changes were observed after miRNA silence, Cells grow normally without obvious changes (Figure 4). We then inhibited hsa-miR-1278 using a synthetic inhibitor in QSG-7701 cells. RT-qPCR verification of hsa-miR-1278 in control and inhibitor groups revealed significant differences between the two groups, which confirmed that hsa-mir-1278 was indeed inhibited (P<0.05) (Figure 5). Western blot was then performed to compare the expression of IL22 and PF4, the potential targets of hsa-miR-1278, in the inhibition cells and the control cells. The results showed that the expression levels PF4 protein are significantly increased in the inhibition group compared to the control group (Figure 6). The miRNA-1278 and PF4 gene expression results were consistent in cellular models with the detection of blood samples from patients. We used biological experiments to confirm that Hsa-miRNA-1278 was unregulated in HE group and therefore reduce the expression of PF4 which was consistent with the database.

**Figure 4.**
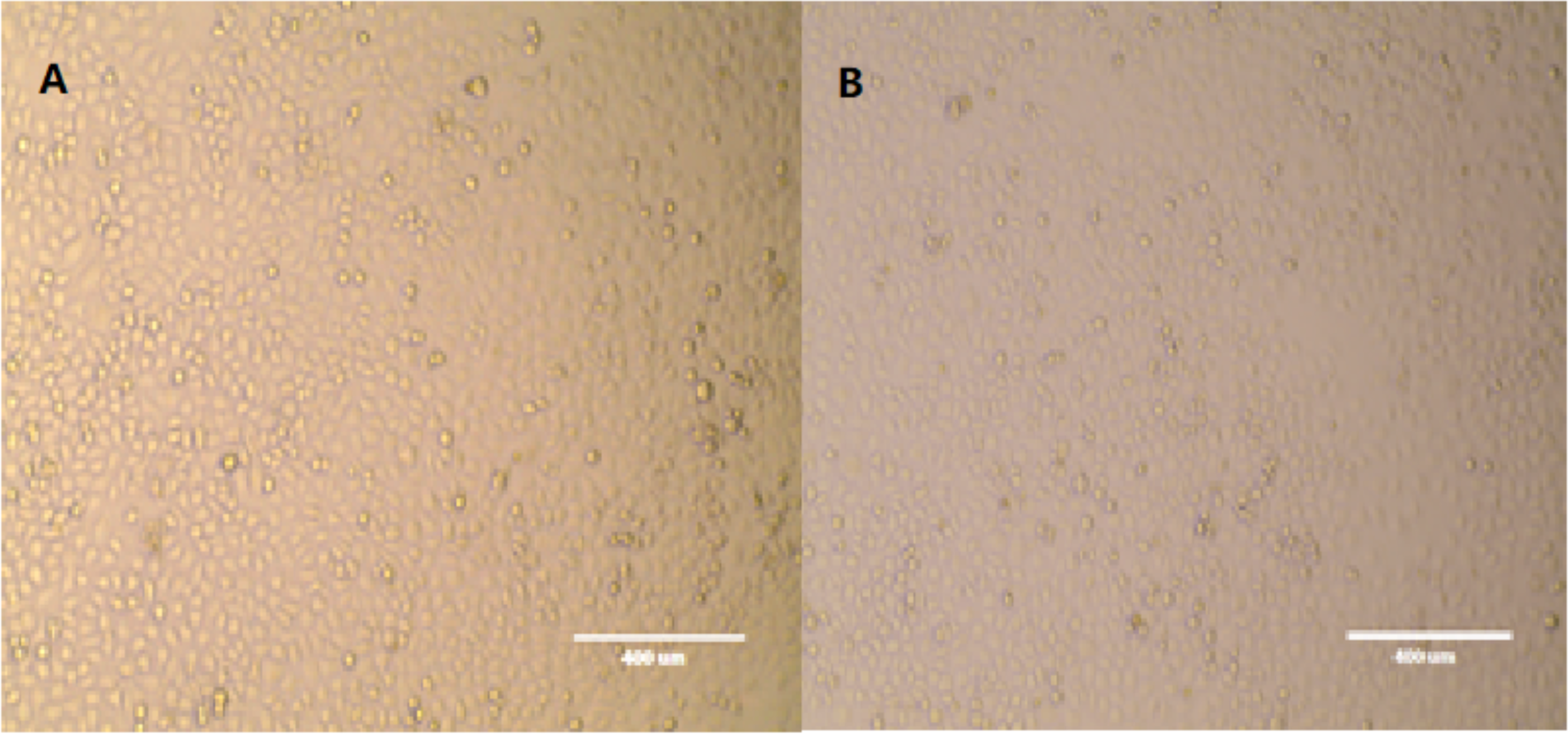
Hsa-miR-1278 silencing in QSG-7701 cells. Normal cell growth and insignificant changes before and after silencing. A: control group; B: Silence group.

**Figure 5.**
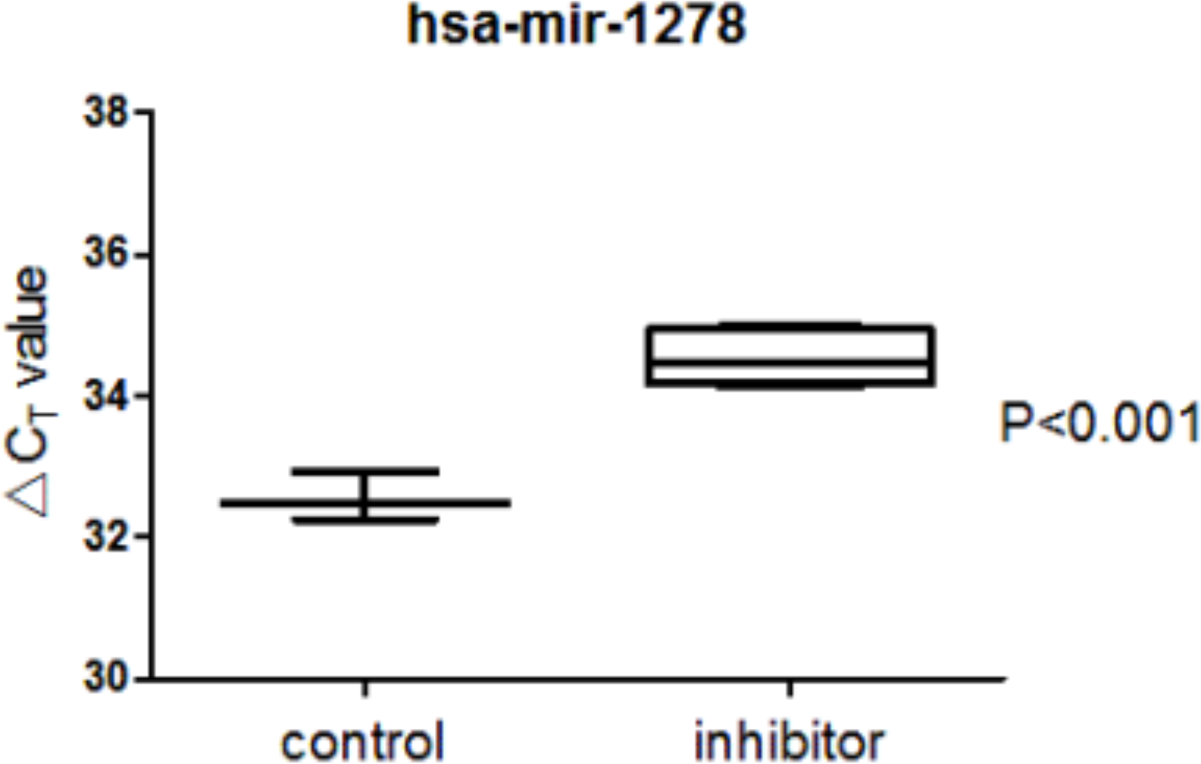
The validation of hsa-miR-1278 in control and inhibitor groups. The inhibitor group was added the synthetic inhibitor to QSG-7701 cells.

**Figure 6.**
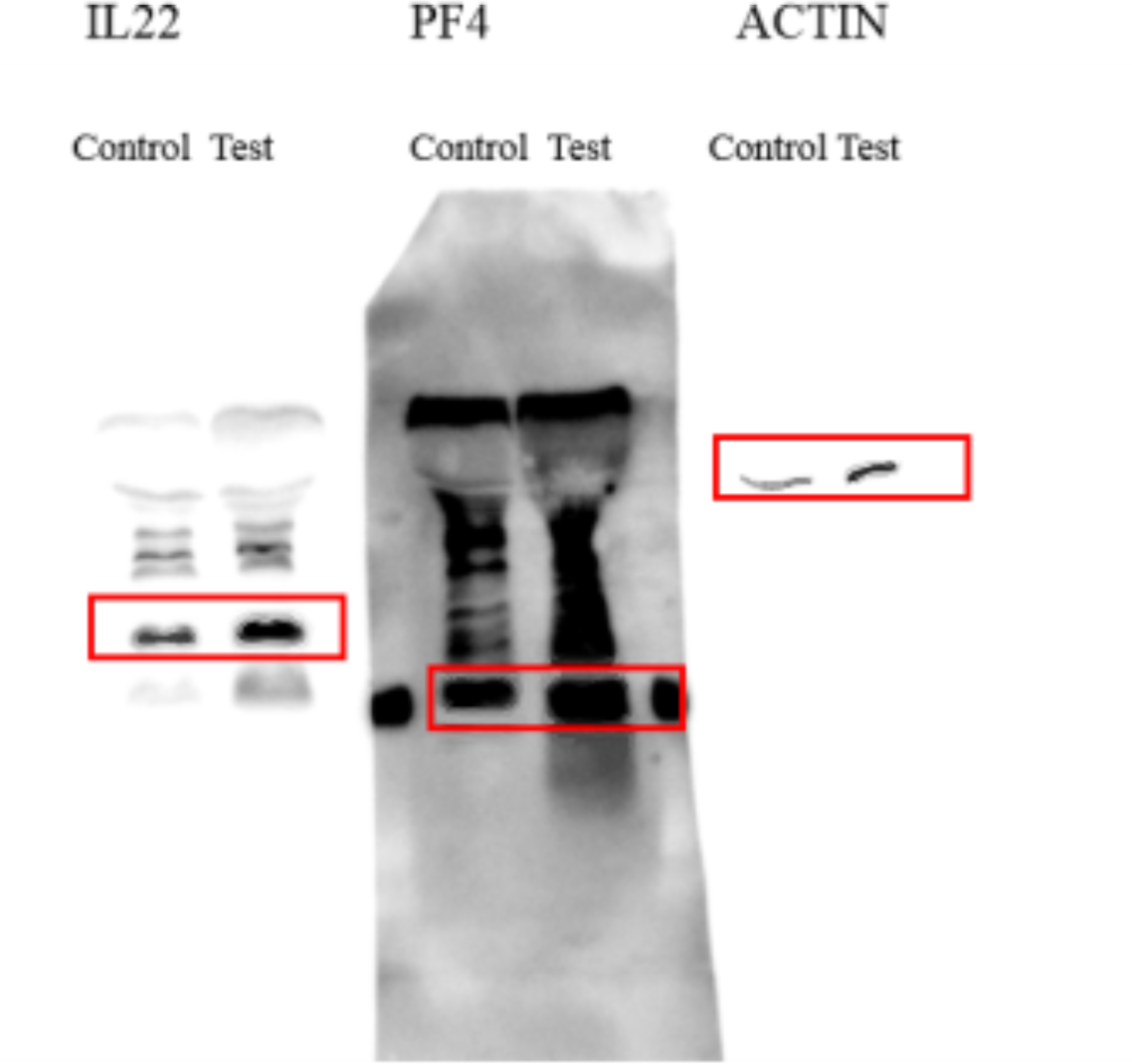
Western blot analysis of IL22 and PF4 in response to hsa-mir-1278 suppression.

### Association adjustment between hsa-miR-1278 and HE

Multivariate adjustment of association between hsa-miR-1278 and hematoma expansion hsa-miR-1278. Differences in clinical characteristics between the HE and non-HE group were analyzed. Univariate analysis indicated that there were significant differences in hsa-miRNA-1278, gender, hypertension history, baseline SBP and baseline ALT between the two groups (Table 2). The association between hsa-miRNA-1278 and hematoma expansion was corrected stepwisely by logistic regression. After adjusted by gender, age, hypertension, baseline SBP and baseline ALT, the expression of hsa-miR-1278 was still significantly up-regulated in the HE group (P<0.05) (Table 3).

**Table 2.** Logistic regression of the relationship between hsa-miRNA-1278 and hematoma expansion

| Factors | *P* value | OR | 95 % CI |  |
| --- | --- | --- | --- | --- |
|  |  |  | Lower | Upper |
| Model 1 * | 0.010 | 0.199 | 0.058 | 0.681 |
| Model 2 † | 0.011 | 0.187 | 0.052 | 0.679 |
| Model 3 ‡ | 0.025 | 0.208 | 0.052 | 0.823 |
| Model 4 \|\| | 0.013 | 0.102 | 0.017 | 0.619 |
| Model 5# | 0.020 | 0.090 | 0.012 | 0.688 |

\*Model 1: univariate analysis of hsa-miRNA-1278
†Model 2: adjusted by gender and age on the basis of Mode 1
‡Mode 3: adjusted by hypertension on the basis of Mode 2
||Mode 4: adjusted by baseline SBP on the basis of Mode 3

## Discussion

Hematoma Expansion has been identified as one of the most important independent determinants of early neurological deterioration and poor clinical outcome after ICH. The pathogenesis influencing the ICH outcome is complex and has been proved to be associated with the pathways of immune response, stress injury repair, and coagulation. The endothelial Wnt/β-catenin pathway (29), some inflammatory factors, such as T cell immunoglobulin and mucin domain-3(TIM-3)(30), Matrix metalloproteinases-9(MMP-9) all contribute to the post-ICH pathogenesis.(31). Among various heated studies, the role of miRNA has been raised continuous attention in recent years. As non-coding RNA family and a powerful regulator of gene expression, microRNA belonging to the non-coding RNA family played an important role in cerebrovascular disease, previously studies revealed that the miR-144(32), miR-155(33), miR-145(34), miR-133b(35) were abnormally expressed in peripheral serum of ICH patients that may serve as the potential regulator of the ICH-induced neuro-inflammation and affect the outcome.

In our study, we found that hsa-miR-1278 was significantly downregulated in the peripheral leukocytes of the ICH patients with HE compared with the control group. After verification via RT-qPCR, we also confirmed that the target gene PF4 was also been remarkably downregulated in the HE group. The miR-1278 was reported to be involved in the pathogenesis of nasopharyngeal carcinoma(36) and cardiovascular disease(37). The hsa-miR-1278 can reduce the expression of IL-22 and CXCL4 by directly binding with the 3’-UTRs of them and therefore downregulate the inflammation response in cardiovascular tissue and apoptosis(38). The hsa-miR-1278 also can promote the human aortic vascular smooth cell proliferation and migration by elevating the DNA methyltransferase 1(DNMT1), which may provide a rational explanation of the process of atherosclerosis vascular disease(38). The hsa-miR-1278 has been also indicated to be involved in the glioblastoma multiforme development through upregulating the LINC00294, a long noncoding RNAs (lncRNAs), to enhance the tumor-suppressing neurofilament medium that ultimately inhibits glioma cell proliferation(39). Based on the above evidences, we believe that the hsa-miR-1278 plays a vital role in the pathobiological way of hematoma expansion in ICH patients.

We performed inhibition experiment and western blotting using QSG-7701 cells (Human hepatocyte line) to confirm that the expression of target gene PF4 was reduced in HE group. The platelet factor 4 is a very abundant platelet α-granule chemokine released during platelet activation. Mature human PF4 is a 7.8 kDa, 70 amino acid protein that exists as a tetramer at physiologic pH and tonicity, which has high affinity for heparin(40), contributing to the pathogenesis of heparin-induced thrombocytopenia and thrombocytopenia syndrome. In vitro studies, PF4 was demonstrated to be procoagulant. The PF4 inhibiting the heparin-dependent acceleration of thrombin inactivation by antithrombin(41) and potentiated platelet aggregation in suboptimal concentrations(42). The recent COVID-19 vaccine-induced thrombocytopenia may provide a vivid process of the PF4-inducing platelet aggregation. The adenovirus-based vaccine ChAdOx1 interacts with PF4 and Coxsackie and Adenovirus receptor (CAR) which expressed in platelet surface and causes the platelet aggregation(43). PF4 also affect hemostasis by inflammatory pathway. After vascular injury, PF4 was found to have various effects on circulating monocytes, preventing apoptosis, boosting phagocytosis and facilitating macrophage differentiation during the inflammation(44). On vascular smooth muscle cell, PF4 cooperates with chemokines like RANTES(45), endothelial E-selectin or endothelial receptors Lipoprotein receptor-related protein(LRP-1)(46) to induce the vascular endothelium proliferation.

In our study, we found a plausible explanation for the formation of HE in ICH patients that the hsa-miR-1278 decreases the expression of platelet factor 4 and therefore upregulate the post-ICH hemostasis and prothrombosis pathways. Therefore, hsa-miR-1278 could be a new target for the treatment of HE in ICH patients. A recent research(47) revealed that the hsa-miR-1278 can be inhibited by fibronectin-1(FN1) in targeted therapy in gastric cancer patients, which can also be a studying-worthy axis to cease HE. We will further explore the possibility of therapeutic strategies and the development of early diagnostic biomarkers.

Potential limitations of our studies should be mentioned. First, all of the patients enrolled in this study were Chinese, so potential selection bias may be inevitable. second, the sample size of this study was relatively small, and further PCR verification with cross-validation is needed in a large population. Third, we select the hepatocyte as the cell line verification, neuron or neurovascular cell shall be selected for further experiments.

## Conclusion

The expression of hsa-miR-1278 was still significantly up-regulated in the hematoma expansion group, and therefore made the hsa-miR-1278 as a novel predictor of ICH prognosis.

## Data Availability

The data that support the findings of this study are available from the corresponding author upon reasonable request.

## Acknowledgments

We thank all of the patients and health care providers who participated in the present study.

## Supplemental material

table 1-2

figure1-6

Reference 1-47

